# Lifestyle interventions for dementia prevention in Low- and Middle-Income Countries: A systematic review

**DOI:** 10.1101/2024.03.01.24303612

**Authors:** Rosario Isabel Espinoza Jeraldo, Sedigheh Zabihi, Claudia Miranda-Castillo, Charles R Marshall, Claudia Cooper

## Abstract

By 2050, two-thirds of people with dementia will live in Low- and Middle-Income Countries (LMICs). Efforts to adapt and test multi-modal prevention interventions focusing on lifestyle changes for people at risk of dementia are being developed predominantly in higher income countries, for people with and without cognitive symptoms. However, there is evidence that needs may differ between these groups. We systematically reviewed Randomized Controlled Trials (RCTs) evaluating non-pharmacological interventions in individuals with Mild Cognitive Impairment (MCI) and Subjective Cognitive Decline (SCC) in LMICs. We analysed study quality using the Mixed Methods Assessment Tool (MMAT), meta-analysed and synthesized evidence. We included 25 RCTs, from six countries (most in China, n=17), involving 1304 participants. Across the 15 studies for which there was sufficient data to meta-analyse, we found significant positive effects on cognitive outcomes favouring interventions [1.49 (standardised mean difference, 95% Confidence Interval= 1.06-1.93)]. There was significant publication bias. Using an a priori standard framework, we classified interventions into exercise, multidomain, and arts/creative expression. Group exercise [1.67, 1.24-2.11, n=8]. and multidomain [1.22, 0.22-2.21, n=5] had replicated evidence of effectiveness. There was insufficient data to meta-analyse the creative arts category. We identified one high quality, multi-modal intervention, which combined Chinese mind-body exercises and health education, reporting a positive outcome. The first robust dementia prevention trials are underway in LMICs, evaluating effectiveness of models developed in higher income countries. We propose greater consideration and investment in development of interventions that account for specific LMIC contexts from the outset, so they are acceptable and used by local services.

**Highlights:**

- There was evidence that group exercise and multimodal interventions were effective for people with memory concerns in LMICs.
- We identified publication bias in meta-analyses; the open science agenda is critical to improving care in LMICs and reducing global inequalities.
- Most included studies were conducted in China, reflecting a need for high-quality evidence from underrepresented regions, including Africa and Latin America.
- We propose greater investment in developing interventions that account for specific LMIC contexts from the outset, so they are acceptable and used by local services.

## 1. Introduction

The number of people living with dementia globally is expected to reach 150 million by 2050, of whom two-thirds will live in the Low- and Middle-Income Countries (LMICs; Martin, A. M., 2006). Risk of dementia is declining in some high-income countries (e.g., Europe and USA), but stable or increasing elsewhere (Naheed et al., 2023). Despite promising results from trials of disease modifying treatments for prodromal and mild Alzheimer’s disease, dementia risk reduction and prevention is the mainstay of national and international policies, including the G8 and World Health Organisation (WHO), to reducing the global dementia burden (Walker & Paddick, 2019).

Dementia is often preceded by a prodromal stage of Subjective Cognitive Complaints (SCC) - where people perceive a decline in their cognitive performance, and/or Mild Cognitive Impairment (MCI), where there is an objective cognitive impairment not meeting criteria for dementia diagnosis (Geda, 2012). In LMICs, prevalence of MCI varies between 6.1% to 30.4%, with approximately 23.8% of people with MCI at risk of Alzheimer’s disease over 3.0 to 5.8 years of follow up (McGrattan et al., 2021; McGrattan et al., 2022). McGrattan highlighted the importance of identifying individuals with cognitive impairment who are at highest risk of dementia in LMICs in order to target risk reduction strategies. A previous systematic review highlighted the moderate effectiveness of lifestyle interventions in reducing cognitive decline, particularly in people with MCI (Whitty et al., 2020).

The FINGER (Finnish Geriatric Intervention Study to Prevent Cognitive Impairment and Disability) trial demonstrated significantly improved cognition in participants aged between 60 and 77 years with vascular risk factors, who received a multi-component intervention (diet, exercise, cognitive training, vascular risk monitoring) relative to a control group over 2 years (Ngandu et al., 2015). These findings are not yet replicated, and no currently available interventions, with proven efficacy, have been demonstrated to reduce dementia cases, or to be scalable to whole populations (Cooper et al., 2021). This intervention was intensive and complex; only 37% of intervention arm participants adhered to at least half of planned activities across four domains (exercise, diet, vascular and cognitive training components). As greater baseline cognitive impairment predicted intervention non-adherence, the authors suggest that additional tailored support for participants at risk of low adherence, which would include those with cognitive concerns, may improve outcomes (Ngandu et al., 2022).

There are increasing efforts to adapt and test multi-modal prevention interventions focusing on lifestyle changes for people at risk of dementia developed in higher income countries, across LMICs. For example, the global “World-Wide FINGERS” initiative is implementing the FINGER multi-modal intervention model in Latin America, Africa and China, in people at risk of dementia with and without cognitive symptoms, though there is evidence needs of these groups may differ (Rosenberg et al., 2020). Economic disparities directly affect access to preventive healthcare, making solutions difficult to design (Prince et al., 2008). Optimism regarding dementia prevention is mitigated by the challenges of large-scale implementation, especially in countries with limited resources (Parra et al., 2019), where socioeconomic contexts often hamper implementation of preventive strategies (Palafox et al., 2016).

Interventions are more likely to be implemented successfully if the contexts in which they are to be used are considered throughout the development process. We systematically reviewed Randomized Controlled Trials (RCTs) evaluating non-pharmacological interventions in individuals with Mild Cognitive Impairment (MCI) and Subjective Cognitive Decline (SCC) in LMICs.

## 2. Methods

We registered this study in the Prospective Register of Systematic Reviews (PROSPERO; registration number: CRD42023403908). We restricted this review to Randomised Controlled Trials (RCTs) testing non-pharmacological dementia secondary prevention strategies, in people with subjective or objective memory concerns.

### 2.1 Search strategy

We systematically searched the electronic databases: MEDLINE (Ovid), PsycINFO (EBSCOhost), Web of Science, LILACS, SciELO and Google Scholar, and reference lists of relevant papers. As national dementia prevention strategies date from after Ngandu et al (2015) demonstrated that non-pharmacological interventions could reduce cognitive decline, we limited our search to after 2010. We search up to 08/03/2023, combining search term strings (English and Spanish) using Boolean operators, selected to mirror the inclusion criteria: 1. Cognitive decline (cognitive impairment OR cognitive decline OR dementia OR demencia OR deterioro cognitivo OR cogni* OR deterioro cognitivo); 2. study design (RCT OR trial OR intervention OR intervención OR ensayo controlado aleatorizado); Country (LMICs OR specific country). The full search strategy is included as Supplementary material (Table 5).

### 2.2 Inclusion and Exclusion criteria

We included RCTs evaluating non-pharmacological interventions aiming to reduce cognitive decline or dementia risk. In line with a previous systematic review (Whitty et al., 2020), we included a broad range of non-pharmacological interventions, where the planned mechanism of action was a change in participants’ lifestyle (diet or exercise), social activities or psychological approaches. We excluded studies of computerised cognitive training interventions, as these were not considered to act through these mechanisms. We also excluded dietary supplements, which we considered pharmacological. We included studies in countries classified as low and middle income by the World Bank at any point between 2010 and 2023; where sample populations were all aged 50 and above and identified as having subjective or objective cognitive concerns (corresponding to a diagnosis of MCI/SCC), however these were measured and identified. We included studies with various control conditions, including treatment as usual, active interventions, or passive controls.

### 2.3 Data extraction

Two researchers, (SZ and RE) independently screened titles and abstracts of identified references using Covidence systematic review management software. Full-text articles from potentially relevant studies were assessed for inclusion. SZ and RE independently extracted relevant data from selected full-text articles. Any disparities in study selection or data extraction were resolved by consulting a third reviewer (CC).

### 2.4 Data Analysis

SZ and RE independently assessed quality of included studies using the RCT items from the Mixed Methods Assessment Tool (MMAT, Hong et al., 2018), which score domain criteria: randomization process, treatment assignment, follow-up, data collection methods, and outcome measurement; we included an additional item similar to Whitty et al (2021), that total sample size for comparisons was 45+; we considered studies that met all six of these criteria to be higher quality, and prioritised their evidence in our synthesis. When we could not find enough information to score an item, we rated it as Uncertain (score = 0). We categorised interventions by type, using previous categorisations as a guide (Klimova & Maresova, 2017; Whitty et al., 2020).

We meta-analysed findings for cognitive outcomes for all studies where we found sufficient information, and then by subtype of intervention. Due to expected heterogeneity and to account for variability within and between studies, we used a random effects model with restricted maximum likelihood (REML) estimator to pool results from included studies using standardized mean differences (SMD) as measure of effect. To assess for heterogeneity, we calculated I^2^, where less than 40% evaluated as not important; 30 to 60% as moderate heterogeneity; and 50 to 90% as substantial heterogeneity (Deeks et al., 2023). Effect sizes were reversed when necessary so higher scores reflected better cognition. We assessed publication bias using funnel plots. Quantitative analysis was performed with R version 4.2.2 using R Studio (Posit team, 2023), and the packages readxl and metafor. For studies that included more than one intervention group, we included the comparison with the control group that demonstrated the greatest positive effect, which in both cases was the more intensive intervention (Lam et al., 2015; Uysal et al., 2023).

## 3. Results

### 3.1 Summary of findings

Figure 1 illustrates the article selection process. We included 25 RCTs, conducted in six countries (China= 17; Brazil= 3; India= 1; Pakistan= 1; Thailand=1; Turkey= 1). 24 studies included people with inclusion criteria equating to Mild Cognitive Impairment, and one participants with SCC (Liu et al., 2023). Intervention durations ranged from one to six months, and all primary outcomes were collected immediately after the intervention periods. In most studies, the primary outcome was global cognition (n=19), and/or cognitive subdomains of memory or executive functions (n = 10). Other primary outcomes included Physical/exercise performance (n=4), Activities of daily living (n=2), emotional status, quality of life and balance confidence (n= 1), and BMI (n= 1). Studies compared interventions to passive (n = 12), active control conditions (n = 11), or both (n=1). No studies reported dementia diagnosis as outcome.

**Figure 1.**
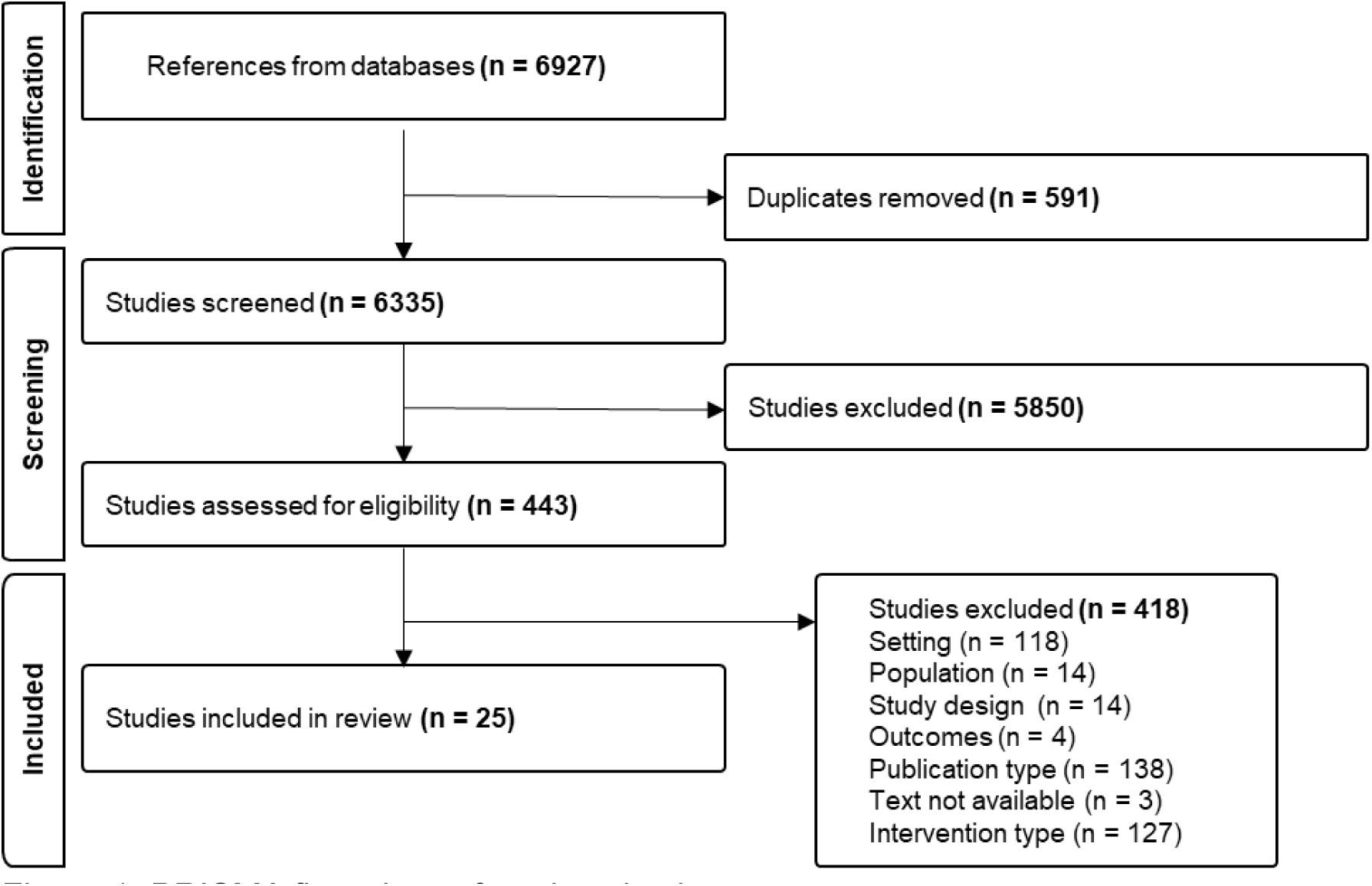
PRISMA flow chart of study selection process

Sufficient data was available to include 15 (60%) of studies in our random effects model analysis evaluating the impact of interventions on cognition outcomes, relative to control conditions (Figure 2a). The estimated overall standardised effect size of the interventions was 1.49 (95% CI:1.06-1.93). The I^2^ estimate, indicated that 46% of the variability in observed results is attributable to between study differences rather than sampling or random errors. The funnel plot suggests publication bias, as we found fewer than expected small studies with negative or null results (Figure 2b). Of the 10 studies we were unable to include, only three attained the highest quality rating.

**Figure 2.**
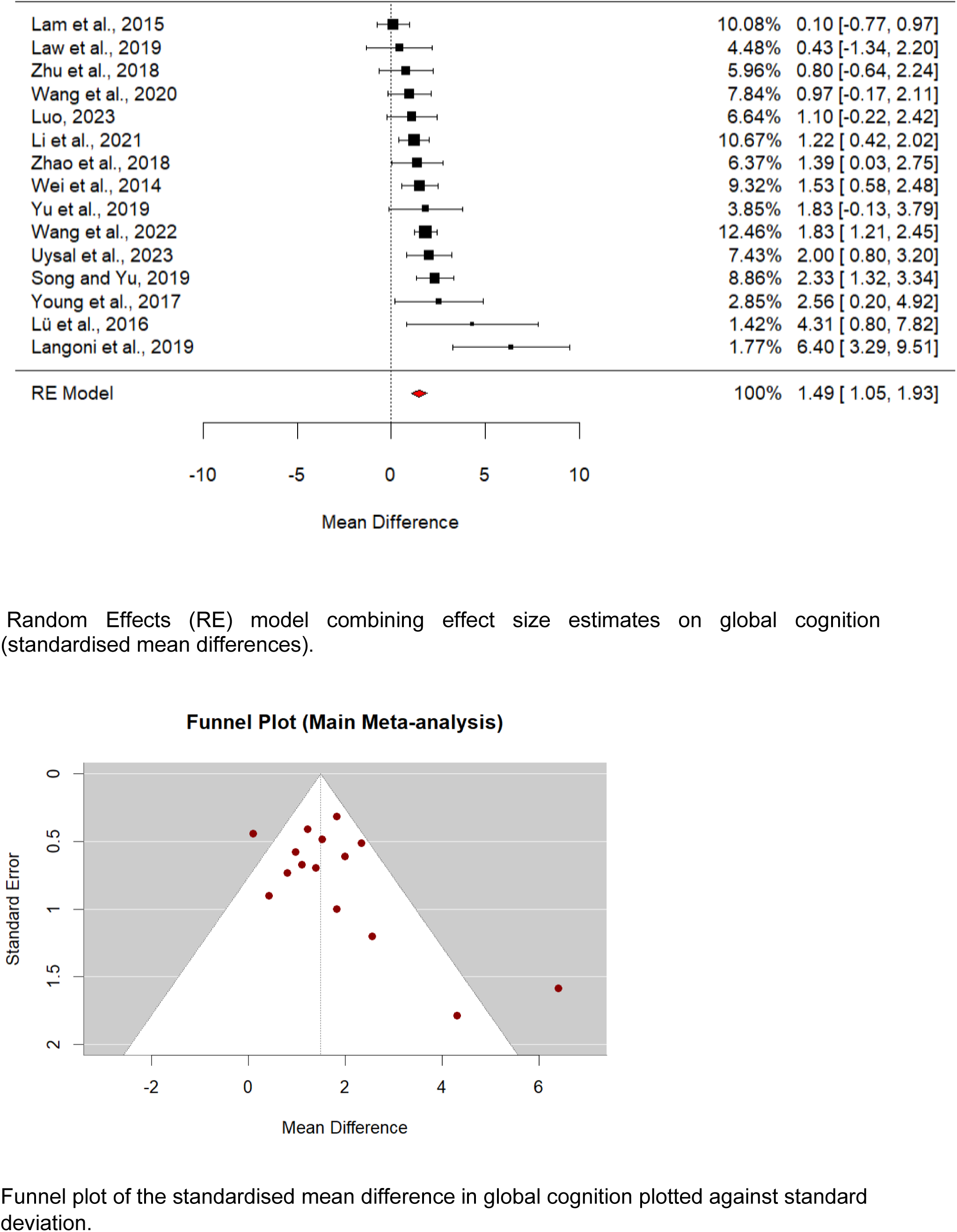
(a) Forest plot demonstrating effects and random effects model for all interventions for which data could be included in the meta-analysis, and (b) funnel pot of included studies.

Table 1 describes quality ratings according to MMAT. 36% (n=9) of studies attained the highest quality rating (all six quality items endorsed), for others five (n = 7), four (n = 5), or three (n = 4) quality items were endorsed. The items relating to randomisation and blinding were most frequently unmet. We classified trials by types of interventions evaluated: exercise only (Table 4; n = 12), multidomain (n = 11; see Table 2 for study characteristics), and arts/creative expression (Table 3; n = 3).

**Table 1:** Summary of studies and reasons for areas of low quality.

| **Author** | **Is randomization appropriately performed?** | **Are the groups comparable at baseline?** | **Are there complete outcome data?** | **Are outcome assessors blinded to the intervention provided?** | **Did the participants adhere to the assigned intervention?** | **Was the sample 45 or more participants?** |
| --- | --- | --- | --- | --- | --- | --- |
| Amjad et al., 2019 | No | Yes | Yes | Uncertain | Yes | No |
| Chatterjee et al., 2022 | Yes | Yes | Uncertain | Uncertain | Yes | No |
| de Oliveira Silva et al., 2019 | Uncertain | Yes | No | Yes | Yes | No |
| Horie et al., 2016 | No | Yes | Yes | Uncertain | No | Yes |
| Kwan et al., 2020 | Yes | Yes | Yes | No | Yes | No |
| Lam et al., 2015 | No | Yes | Yes | Yes | Yes | Yes |
| Langoni, 2019 | Uncertain | Yes | Yes | yes | Yes | Yes |
| Law et al., 2019 | Yes | Yes | Yes | Yes | Yes | No |
| Li et al., 2021 | Uncertain | Yes | Uncertain | Yes | Uncertain | Yes |
| Lin et al., 2022 | Yes | Yes | No | Yes | Yes | Yes |
| Liu et al., 2023 | Yes | No | Yes | Yes | Yes | Yes |
| Lü et al., 2016 | Yes | Yes | Yes | Yes | Yes | Yes |
| Luo et al., 2023 | Yes | Yes | Yes | Yes | Uncertain | Yes |
| Song & Yu, 2019 | Yes | Yes | Yes | Yes | Yes | Yes |
| Sungkart et al., 2018 | Yes | Yes | Yes | Yes | Yes | Yes |
| Tao et al., 2019 | Yes | Yes | Yes | Yes | Yes | Yes |
| Uysal et al., 2023 | Yes | Yes | Yes | Yes | Yes | No |
| Wang et al., 2020 | Yes | Yes | Yes | Yes | Yes | Yes |
| Wang et al., 2022 | Yes | Yes | Yes | Yes | Yes | Yes |
| Wei et al., 2014 | No | Yes | Yes | No | Yes | Yes |
| Young et al., 2017 | No | Yes | Yes | Yes | Yes | No |
| Yu et al., 2019 | Yes | Yes | Yes | Uncertain | Uncertain | Yes |
| Zhu et al., 2018 | Yes | Yes | Yes | Yes | Yes | Yes |
| Zhao et al., 2018 | Yes | Yes | Yes | Yes | Yes | Yes |
| Griffiths et al, 2020 | Yes | Yes | Yes | Yes | Yes | Yes |

**Table 2:**
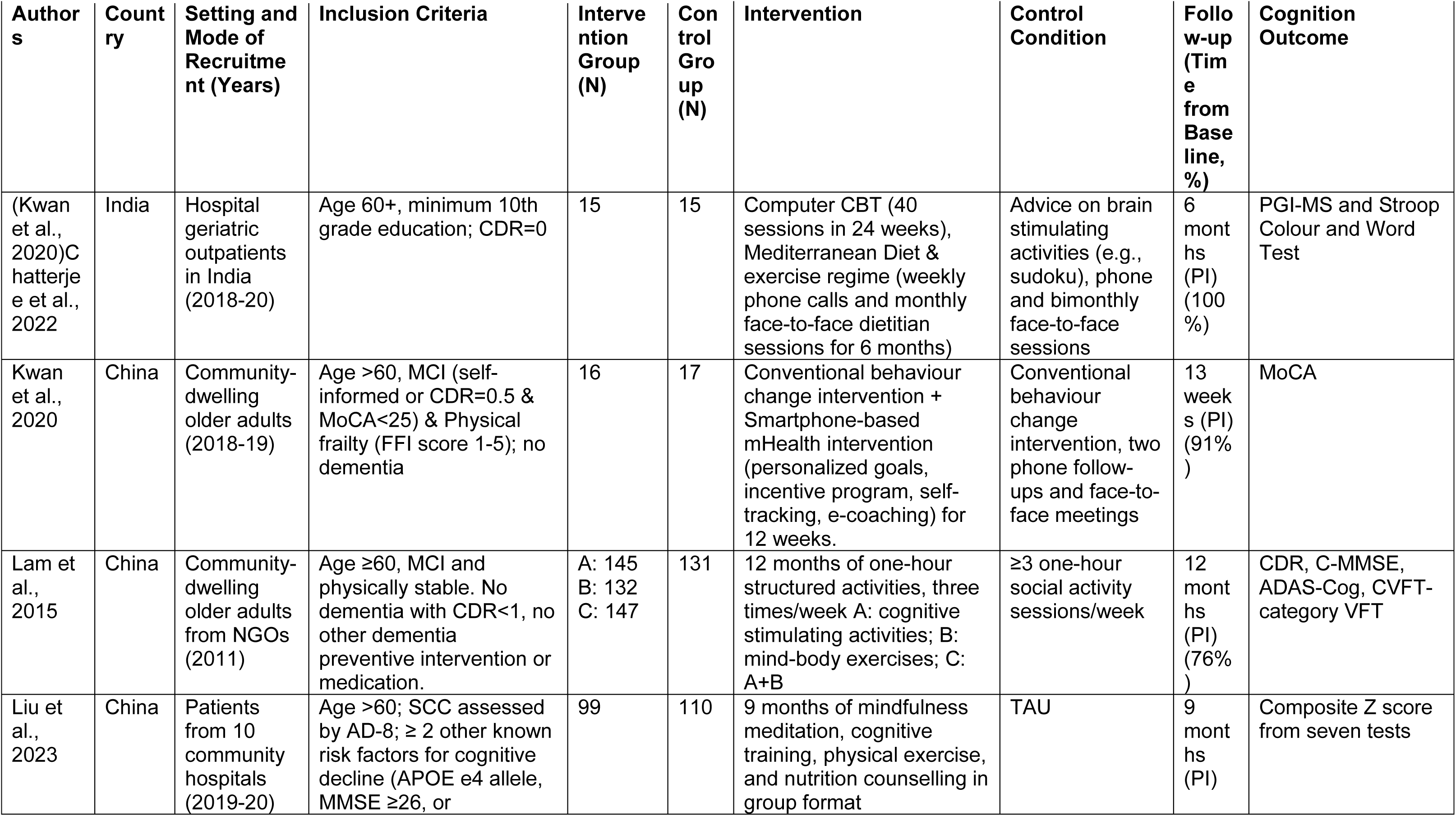
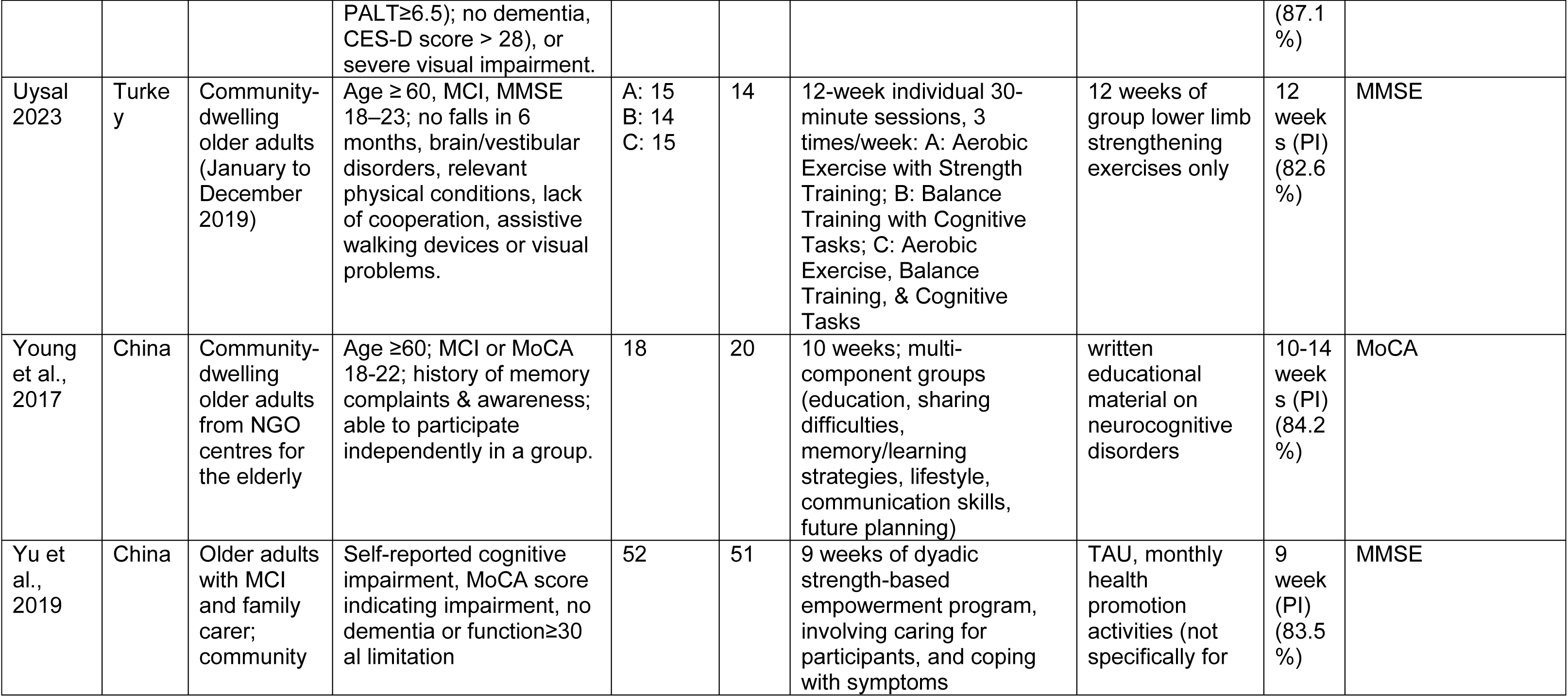
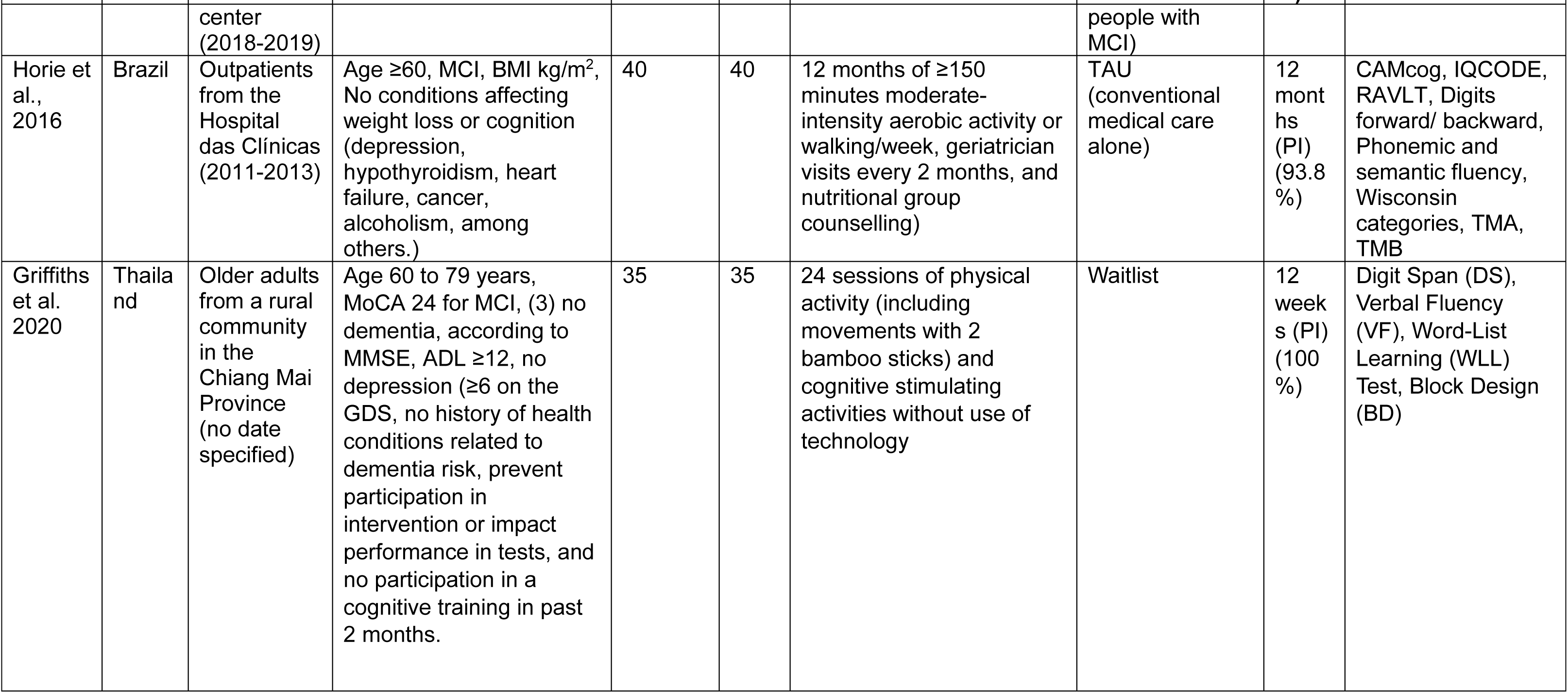
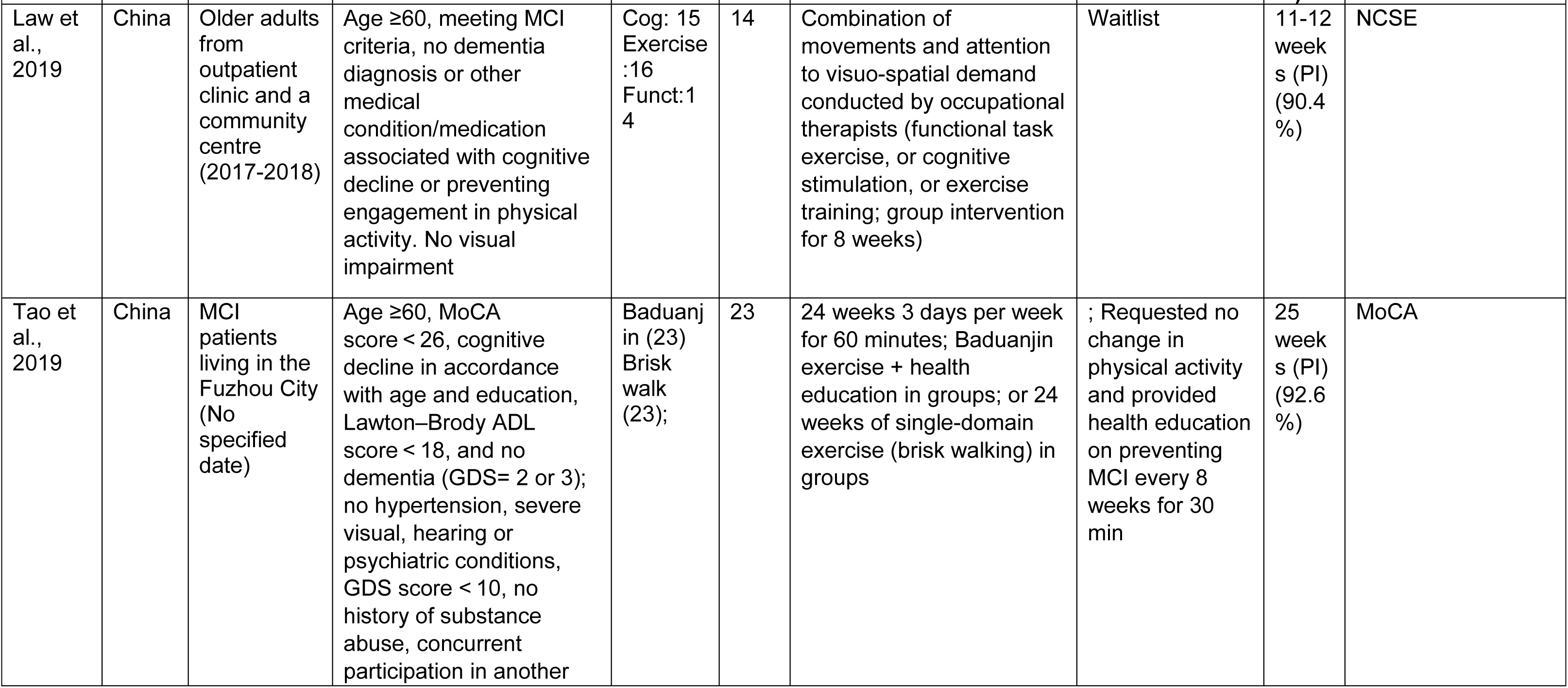
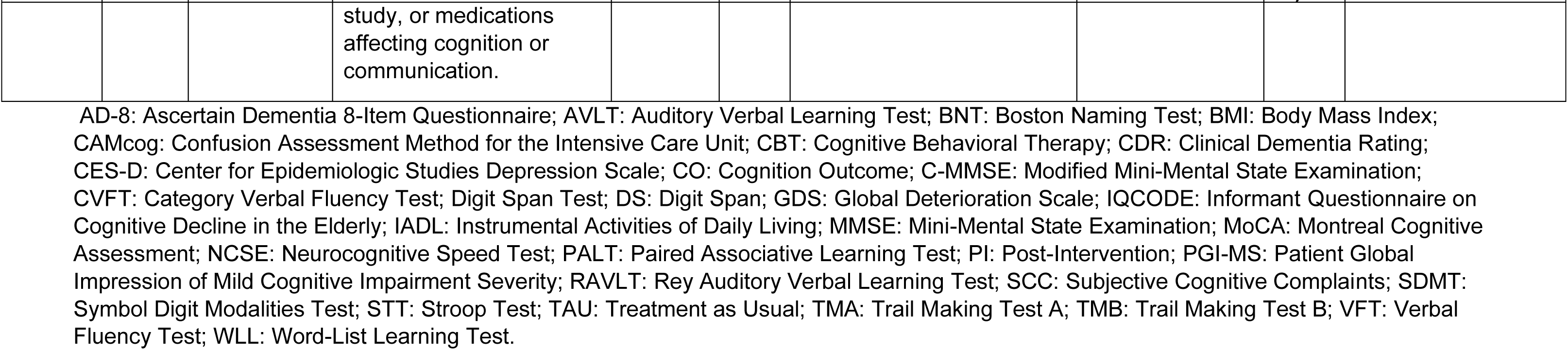
Characteristics of trials evaluating multimodal interventions.

| Author<br>s | Count<br>ry | Setting and<br>Mode of<br>Recruitme<br>nt (Years) | Inclusion Criteria | Interve<br>ntion<br>Group<br>(N) | Con<br>trol<br>Gro<br>up<br>(N) | Intervention | Control<br>Condition | Follo<br>w-up<br>(Tim<br>e<br>from<br>Base<br>line,<br>%) | Cognition<br>Outcome |
| --- | --- | --- | --- | --- | --- | --- | --- | --- | --- |
| (Kwan et al., 2020)Chatterjee et al., 2022 | India | Hospital geriatric outpatients in India (2018-20) | Age 60+, minimum 10th grade education; CDR=0 | 15 | 15 | Computer CBT (40 sessions in 24 weeks), Mediterranean Diet & exercise regime (weekly phone calls and monthly face-to-face dietitian sessions for 6 months) | Advice on brain stimulating activities (e.g., sudoku), phone and bimonthly face-to-face sessions | 6 months (PI) (100%) | PGI-MS and Stroop Colour and Word Test |
| Kwan et al., 2020 | China | Community-dwelling older adults (2018-19) | Age >60, MCI (self-informed or CDR=0.5 & MoCA<25) & Physical frailty (FFI score 1-5); no dementia | 16 | 17 | Conventional behaviour change intervention + Smartphone-based mHealth intervention (personalized goals, incentive program, self-tracking, e-coaching) for 12 weeks. | Conventional behaviour change intervention, two phone follow-ups and face-to-face meetings | 13 weeks (PI) (91%) | MoCA |
| Lam et al., 2015 | China | Community-dwelling older adults from NGOs (2011) | Age ≥60, MCI and physically stable. No dementia with CDR<1, no other dementia preventive intervention or medication. | A: 145<br>B: 132<br>C: 147 | 131 | 12 months of one-hour structured activities, three times/week A: cognitive stimulating activities; B: mind-body exercises; C: A+B | ≥3 one-hour social activity sessions/week | 12 months (PI) (76%) | CDR, C-MMSE, ADAS-Cog, CVFT-category VFT |
| Liu et al., 2023 | China | Patients from 10 community hospitals (2019-20) | Age >60; SCC assessed by AD-8; ≥ 2 other known risk factors for cognitive decline (APOE e4 allele, MMSE ≥26, or | 99 | 110 | 9 months of mindfulness meditation, cognitive training, physical exercise, and nutrition counselling in group format | TAU | 9 months (PI) | Composite Z score from seven tests |
|  |  |  | PALT≥6.5); no dementia, CES-D score > 28), or severe visual impairment. |  |  |  |  | (87.1 %) |  |
| Uysal<br>2023 | Turke<br>y | Community-dwelling<br>older adults<br>(January to<br>December<br>2019) | Age ≥ 60, MCI, MMSE 18–23; no falls in 6 months, brain/vestibular disorders, relevant physical conditions, lack of cooperation, assistive walking devices or visual problems. | A: 15<br>B: 14<br>C: 15 | 14 | 12-week individual 30-minute sessions, 3 times/week: A: Aerobic Exercise with Strength Training; B: Balance Training with Cognitive Tasks; C: Aerobic Exercise, Balance Training, & Cognitive Tasks | 12 weeks of group lower limb strengthening exercises only | 12 week<br>s (PI)<br>(82.6 %) | MMSE |
| Young<br>et al.,<br>2017 | China | Community-dwelling<br>older adults<br>from NGO<br>centres for<br>the elderly | Age ≥60; MCI or MoCA 18-22; history of memory complaints & awareness; able to participate independently in a group. | 18 | 20 | 10 weeks; multi-component groups (education, sharing difficulties, memory/learning strategies, lifestyle, communication skills, future planning) | written educational material on neurocognitive disorders | 10-14 week<br>s (PI)<br>(84.2 %) | MoCA |
| Yu et<br>al.,<br>2019 | China | Older adults<br>with MCI<br>and family<br>carer;<br>community | Self-reported cognitive impairment, MoCA score indicating impairment, no dementia or functional limitation | 52 | 51 | 9 weeks of dyadic strength-based empowerment program, involving caring for participants, and coping with symptoms | TAU, monthly health promotion activities (not specifically for | 9 week<br>(PI)<br>(83.5 %) | MMSE |

| Author<br>s | Count<br>ry | Setting and<br>Mode of<br>Recruitme<br>nt (Years) | Inclusion Criteria | Interve<br>nion<br>Group<br>(N) | Con<br>trol<br>Gro<br>up<br>(N) | Intervention | Control<br>Condition | Follo<br>w-up<br>(Tim<br>e<br>from<br>Base<br>line,<br>%) | Cognition<br>Outcome |
| --- | --- | --- | --- | --- | --- | --- | --- | --- | --- |
|  |  | center<br>(2018-2019) |  |  |  |  | people with<br>MCI) |  |  |
| Horie et<br>al.,<br>2016 | Brazil | Outpatients<br>from the<br>Hospital<br>das Clínicas<br>(2011-2013) | Age ≥60, MCI, BMI kg/m<sup>2</sup>,<br>No conditions affecting<br>weight loss or cognition<br>(depression,<br>hypothyroidism, heart<br>failure, cancer,<br>alcoholism, among<br>others.) | 40 | 40 | 12 months of ≥150<br>minutes moderate-intensity aerobic activity or<br>walking/week, geriatrician<br>visits every 2 months, and<br>nutritional group<br>counselling) | TAU<br>(conventional<br>medical care<br>alone) | 12<br>mont<br>hs<br>(PI)<br>(93.8<br>%) | CAMcog, IQCODE,<br>RAVLT, Digits<br>forward/ backward,<br>Phonemic and<br>semantic fluency,<br>Wisconsin<br>categories, TMA,<br>TMB |
| Griffiths<br>et al.<br>2020 | Thaila<br>nd | Older adults<br>from a rural<br>community<br>in the<br>Chiang Mai<br>Province<br>(no date<br>specified) | Age 60 to 79 years,<br>MoCA 24 for MCI, (3) no<br>dementia, according to<br>MMSE, ADL ≥12, no<br>depression (≥6 on the<br>GDS, no history of health<br>conditions related to<br>dementia risk, prevent<br>participation in<br>intervention or impact<br>performance in tests, and<br>no participation in a<br>cognitive training in past<br>2 months. | 35 | 35 | 24 sessions of physical<br>activity (including<br>movements with 2<br>bamboo sticks) and<br>cognitive stimulating<br>activities without use of<br>technology | Waitlist | 12<br>week<br>s (PI)<br>(100<br>%) | Digit Span (DS),<br>Verbal Fluency<br>(VF), Word-List<br>Learning (WLL)<br>Test, Block Design<br>(BD) |
| Law et<br>al.,<br>2019 | China | Older adults<br>from<br>outpatient<br>clinic and a<br>community<br>centre<br>(2017-2018) | Age ≥60, meeting MCI<br>criteria, no dementia<br>diagnosis or other<br>medical<br>condition/medication<br>associated with cognitive<br>decline or preventing<br>engagement in physical<br>activity. No visual<br>impairment | Cog: 15<br>Exercise<br>:16<br>Funct:1<br>4 | 14 | Combination of<br>movements and attention<br>to visuo-spatial demand<br>conducted by occupational<br>therapists (functional task<br>exercise, or cognitive<br>stimulation, or exercise<br>training; group intervention<br>for 8 weeks) | Waitlist | 11-12<br>week<br>s (PI)<br>(90.4<br>%) | NCSE |
| Tao et<br>al.,<br>2019 | China | MCI<br>patients<br>living in the<br>Fuzhou City<br>(No<br>specified<br>date) | Age ≥60, MoCA<br>score < 26, cognitive<br>decline in accordance<br>with age and education,<br>Lawton–Brody ADL<br>score < 18, and no<br>dementia (GDS= 2 or 3);<br>no hypertension, severe<br>visual, hearing or<br>psychiatric conditions,<br>GDS score < 10, no<br>history of substance<br>abuse, concurrent<br>participation in another study, or medications<br>affecting cognition or<br>communication. | Baduanj<br>in (23)<br>Brisk<br>walk<br>(23); | 23 | 24 weeks 3 days per week<br>for 60 minutes; Baduanjin<br>exercise + health<br>education in groups; or 24<br>weeks of single-domain<br>exercise (brisk walking) in<br>groups | ; Requested no<br>change in<br>physical activity<br>and provided<br>health education<br>on preventing<br>MCI every 8<br>weeks for 30<br>min | 25<br>week<br>s (PI)<br>(92.6<br>%) | MoCA |

AD-8: Ascertain Dementia 8-Item Questionnaire; AVLT: Auditory Verbal Learning Test; BNT: Boston Naming Test; BMI: Body Mass Index; CAMcog: Confusion Assessment Method for the Intensive Care Unit; CBT: Cognitive Behavioral Therapy; CDR: Clinical Dementia Rating; CES-D: Center for Epidemiologic Studies Depression Scale; CO: Cognition Outcome; C-MMSE: Modified Mini-Mental State Examination; CVFT: Category Verbal Fluency Test; Digit Span Test; DS: Digit Span; GDS: Global Deterioration Scale; IQCODE: Informant Questionnaire on Cognitive Decline in the Elderly; IADL: Instrumental Activities of Daily Living; MMSE: Mini-Mental State Examination; MoCA: Montreal Cognitive Assessment; NCSE: Neurocognitive Speed Test; PALT: Paired Associative Learning Test; PI: Post-Intervention; PGI-MS: Patient Global Impression of Mild Cognitive Impairment Severity; RAVLT: Rey Auditory Verbal Learning Test; SCC: Subjective Cognitive Complaints; SDMT: Symbol Digit Modalities Test; STT: Stroop Test; TAU: Treatment as Usual; TMA: Trail Making Test A; TMB: Trail Making Test B; VFT: Verbal Fluency Test; WLL: Word-List Learning Test.

**Table 3:**
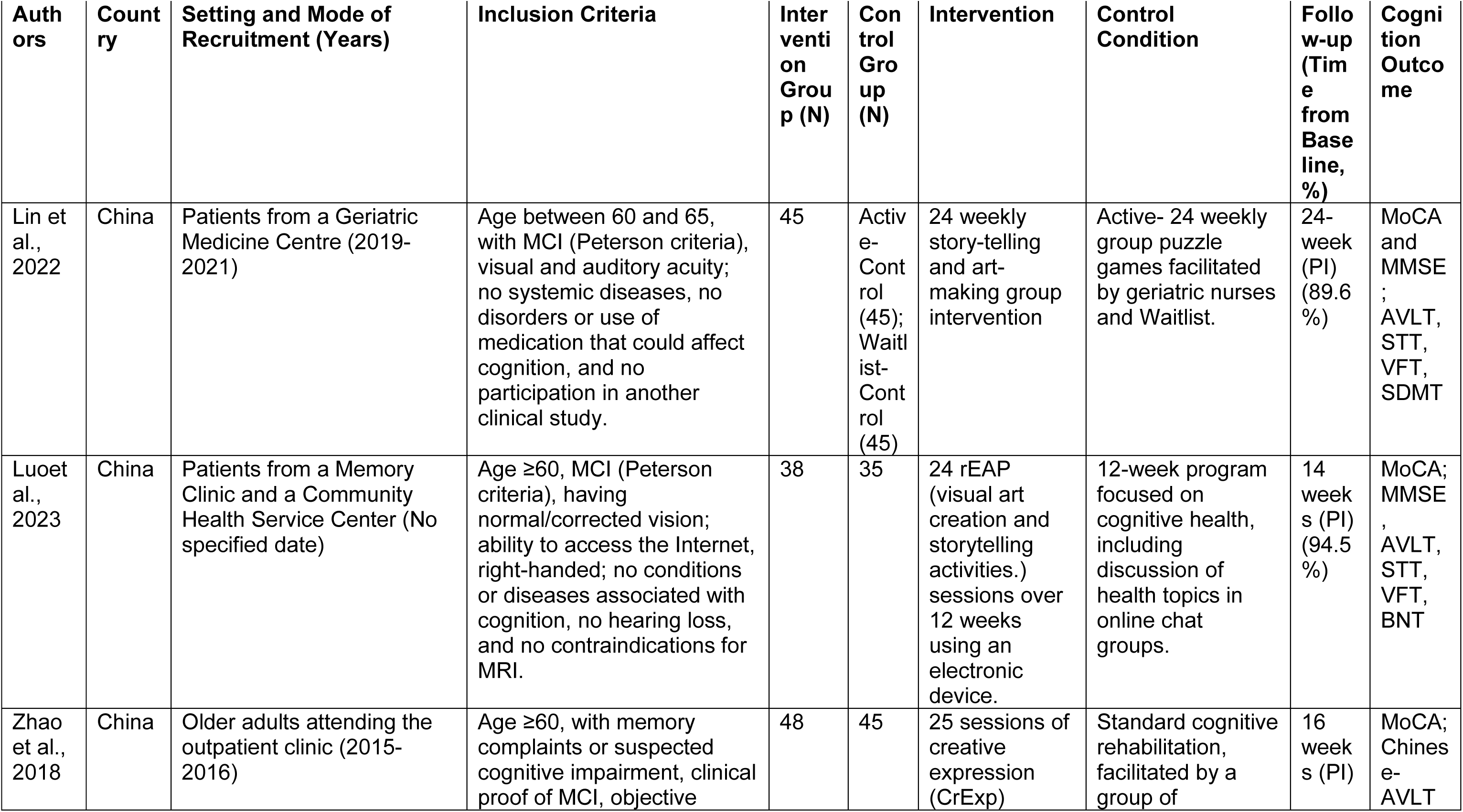
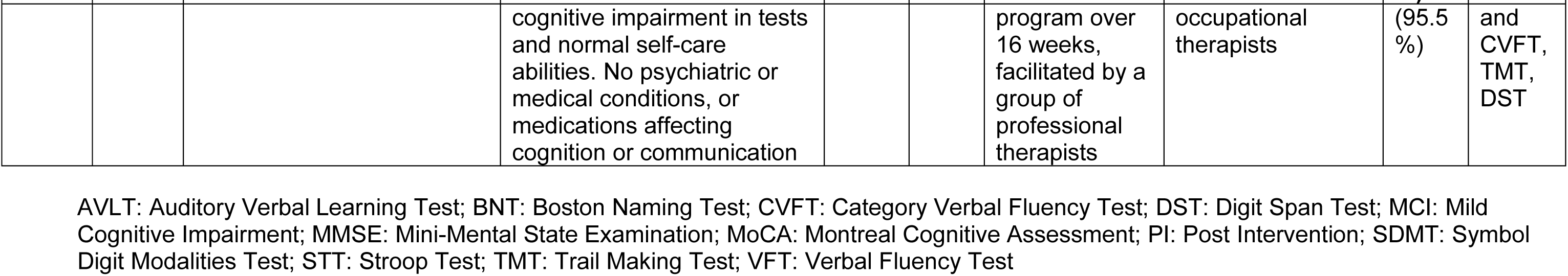
Characteristics of trials evaluating arts-based interventions.

**Table 4.** Characteristics of trials evaluating exercise only interventions.

| Authors | Country | Setting and Mode of Recruitment (Years) | Inclusion Criteria | Intervention Group (N) | Control Group (N) | Intervention | Control Condition | Follow-up (Time from Baseline, %) | Cognitive Outcome |
| --- | --- | --- | --- | --- | --- | --- | --- | --- | --- |
| Amjad et al., 2019 | Pakistan | Patients from a hospital physical therapy department | Patients with MCI; MMSE & MoCA <25; able to perform aerobic exercises; adequate vision and hearing; able to consent; no brain, cardiac, neurological, psychiatric, renal, or systemic conditions; no drug use or sleep disturbance. | 19 | 21 | Aerobic exercise group three days per week for six weeks | Advice on home-based gentle movements and stretching 3x a week for 6 weeks. | 6 weeks (PI) (82.5%) | MoCA and MMSE |
| de Oliveira Silva et al., 2019 | Brazil | Patients from the Center of Alzheimer's, Federal University of Rio de Janeiro (2016-2018) | Age ≥60, MCI, BMI ≥ 30 kg/m<sup>2</sup>; Absence of conditions interfering with weight loss or cognition, and recent use of specific medications (anti-obesity drugs, benzodiazepines, neuroleptics, or estrogen in past 2 months). | 7 | 12 | 60 min. sessions twice a week for 12 weeks, including balance, aerobic, strength, and stretching, supervised by physiotherapists. | Treatment as usual | 3 months (PI) (100%) | MMSE, Clock drawing test, Verbal Fluency, Stroop test |
| Langoni, 2019 | Brazil | Older users of 4 primary care units in Porto Alegre/RS, Brazil (October 2015-March 2017) | Age ≥60, MCI, sedentary, independent in activities of daily living; no severe psychiatric/neurological illnesses or physical conditions impacting participation or cognition, acetylcholinesterase inhibitor, communication deficits, concurrent study participation, recent physiotherapy, uncontrolled hypertension. | 30 | 30 | Twice-weekly group exercise sessions for 24 weeks, conducted by a qualified physical therapist (approximately 60 minutes each) | Control group was asked not to initiate any physical or cognitive activity during the intervention period. Regular telephone contact to monitor this | 24 week (PI) (86.6%) | MMSE |
| Li et al., 2021 | China | Older adults from the Wang Yuehu community (No specified date) | Age $\geq 60$, with no hearing or visual issues, MoCA scores according to educational levels, meeting MCI or subjective memory decline criteria, unaffected daily life, and no dementia. No serious physical disease or regular exercise in 6 months. | 45 | 45 | Multi-component group exercise training program, 5 times per week for 6 months | Community health instruction (i.e., receiving lessons lasting 1 hour every month including how to keep fit) at 3 and 6 months | 3 months (PI) (93.3%) | MMSE and MoCA |
| Lü et al., 2016 | China | Community-dwelling older adults (No specified date) | Age $\geq 65$, memory complaints, MMSE $\geq 24$, MoCA $< 26$, ADL scale $< 26$ and no dementia or conditions associated with cognition and physical performance, not participating in any regular physical activities. | 22 | 23 | 12 weeks; three times a week, single domain (momentum-based dumbbell training) | Maintain a regular lifestyle routine without starting any new exercise activities | 12 weeks (PI) (93.3%) | ADAS-Cog; TMT-B and Digit Span Chinese version |
| Song and Yu, 2019 | China | Community-dwelling adults from two community healthcare centers (2017-2018) | Age $\geq 60$, MCI (MoCA-C between 19 and 26), no conditions contraindicated for exercise training, no severe neurological disorders or use of medication that greatly impair cognitive function. | 60 | 60 | 16-week aerobic stepping exercise program with three 60-minute group training sessions per week | Health education program delivered by a general practitioner for 16 weeks | 16 weeks (PI) (96.6%) | MoCA |
| Sungkarat et al., 2018 | Thailand | Amnesic multiple-domain MCI recruited from the local community | MMSE $\geq 24$, MoCA $< 26$, no use of cognitive impairment medications, absence of neurological conditions like Parkinson's disease or stroke, no depression, uncorrected vision and/or hearing issues, and no | 33 | 33 | Practice of Tai Chi for 6 months (9 group sessions). After that, home practice three times per week for | a 1-hour presentation on cognitive impairment and falls plus information booklet for prevention. | 6 months (PI) (84.4%) | LM delayed recall, Block Design Test, |
|  |  | (No specified date) | conditions preventing exercise and those already engaged in regular exercise. |  |  | the remaining 6 months. |  |  | Digit Span and Trail Making Test |
| Wang et al., 2022 | China | Community-dwelling older persons with MCI (No specified date) | Age $\geq 60$, MCI with MMSE scores depending on education level); no dementia, severe neurological or psychiatric disorders, no significant hearing or visual impairments, history of substance abuse, physical conditions hindering participation, or use of cognitive-affecting medications. | 117 | 115 | Individual exercise intervention consisting of 30 minutes of finger exercises twice daily for 30 days | Waiting list to receive the same finger exercise after the end of the study | 30 days (PI) (98.3%) | MMSE |
| Wang et al., 2020 | China | Community-dwelling adults from community senior and healthcare centers (June 2018-October 2019) | Age $\geq 60$ years old, MCI with MMSE 25–30 or MoCA of $\leq 26$; intact activities of daily living. No regular exercise, no significant health conditions, impairments, or medicine use that could interfere with cognitive performance or with engaging in physical exercise, and no participation in other studies. | 57 | 54 | Structured limbs-exercise program for 12 weeks, followed by 12 weeks of unsupervised at home. Supervised by physiotherapists with 5-year experience. | Waiting list to receive the same intervention after follow-up, attending to a 5-minute health promotion class twice over the 12 weeks | 12 weeks (PI) (93.51%) | MoCA |
| Wei et al., 2014 | China | Older adults with MCI in a nursing home (October | 60 to 75 years old, $\geq 3$ months subjective or objective cognitive impairment, MMSE $\leq 26$ points, GDS 2-3, ADL Score $\leq 18$ points, HIS $\leq 4$ | 30 | 30 | 6 months of exercise group intervention | Treatment as usual | 6 months (PI) (100%) | MMSE |

| Aut<br>hors | Cou<br>ntry | Setting and<br>Mode of<br>Recruitment<br>(Years) | Inclusion Criteria | Inter<br>vent<br>ion<br>Gro<br>up<br>(N) | Con<br>trol<br>Gro<br>up<br>(N) | Intervention | Control Condition | Follow-up<br>(Time<br>from<br>Baseli<br>ne, %) | Cognitio<br>n<br>Outcom<br>e |
| --- | --- | --- | --- | --- | --- | --- | --- | --- | --- |
|  |  | 2011-August<br>2012) | points; normal or corrected-to-<br>normal hearing and vision; SRDS<br>≤53, no history of drug use or<br>movement disorder. |  |  |  |  |  |  |
| Zhu<br>et<br>al.,<br>2018 | Chin<br>a | Patients from<br>dementia<br>clinic or<br>recruited<br>through<br>radio/newspa<br>per<br>recruitment<br>(2014-2015) | Age between 50 and 85 years, MCI<br>with reported memory problems for<br>at least 3 months and MMSE score<br>≥25 and a MoCA score ≤26. | 29 | 31 | 35-minute dance<br>session 3 times a<br>week for 3 months | Treatment as Usual | 3<br>months<br>(PI)<br>(96.6%) | MoCA;<br>Wechler<br>Memory<br>Scale,<br>Symbol<br>Digit<br>Modalitie<br>s Test,<br>Trail<br>Making<br>Test,<br>Digit<br>Span |

ADAS-Cog: Alzheimer's Disease Assessment Scale - Cognitive Subscale; ADL: Activities of Daily Living; BMI: Body Mass Index; BDT: Block Design Test; GDS: Geriatric Depression Scale; HIS: Hachinski Ischemic Score; LM: Logical Memory; MCI: Mild Cognitive Impairment; MMSE: Mini-Mental State Examination; MoCA: Montreal Cognitive Assessment; SDMT: Symbol Digit Modalities Test; SRDS: Digit Span; TMT-B: Trail Making Test B; WMS: Wechsler Memory Scale

### 3.2 Exercise interventions (Figure 3)

#### 3.2.1 Studies rated as highest quality (n=6)

Interventions evaluated in these studies ranged from one to six months in duration, all incorporated social elements. All studies were set in China, except for Sungkarat and collaborators (2018) set in Thailand. Of the six studies rated as high quality, in four the primary outcome favoured the intervention. Lü and colleagues (2016) evaluated three months of thrice-weekly momentum-based dumbbell group training in a community centre, delivered by previously trained students (adherence rate: 93.3%); a significant difference in the primary cognitive outcome (Alzheimer’s Disease Assessment Scale – Cognitive) favoured the 60 minute intervention. Song and Yu (2019) implemented a four-month group based aerobic stepping exercise program, which incorporated elements such as goal setting and self-monitoring and sought to encourage movements that could be easily integrated into daily activities. The intervention was facilitated by two registered nurses (adherence rate: 73.1%). Wang and colleagues (2022) tested a 30-minute finger exercise session twice daily for one month (adherence rate 98.3%). Participants were taught the exercise routine and encouraged to practice it daily. Although the intervention was individual, participants in this group had a telephone check-up twice every morning and evening with a researcher to ensure adherence while control participants did not. Sungkarat et al. (2018), the only high-quality study we were unable to include in the meta-analysis, nine Tai Chi group training sessions, followed by 6 months home practice with weekly interactions with a researcher (adherence rate: 83.6%) improved memory and executive functions in the intervention group, relative to the control group.

**Figure 3:**
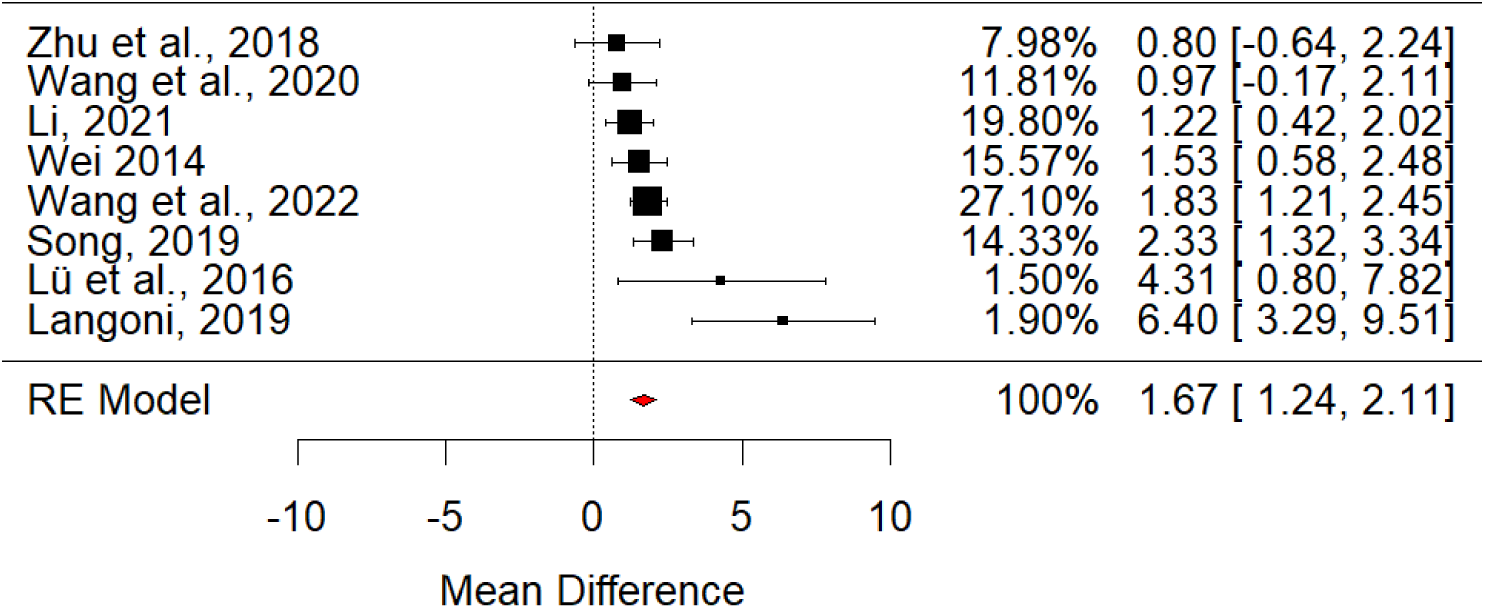
Meta analysis of exercise interventions.

Two high quality exercise studies, both included in our meta-analysis (Figure 3), did not find in favour of the intervention. Zhu and collaborators (2018) tested a 35-minute session (thrice weekly for three months) facilitated by a dance instructor; the study, which lacked statistical power, did not find significant differences between groups (adherence rate; 88.9%). Wang et al. (2020) evaluated three months of a weekly, structured, 60 minutes exercise group; with an experienced physiotherapist, followed by three months of unsupervised home sessions (adherence rate: 93.5%). The cognition outcome favoured the intervention at three (mean difference = 1.20, 95% CI [0.354, 2.054], p = 0.006) and six months follow up, though this comparison did not attain significance in our meta-analysis, which additionally controlled for baseline cognition.

#### 3.2.2 Studies rated as lower quality (n=5)

The three studies rated as lower quality that we included in our meta-analysis evaluated six-month, group interventions, and all reported findings favouring the intervention. These included a twice-weekly programme led by a trained physical therapist in Brazil (Langoni et al., 2019), a multi-component programme conducted in China held five times per week (Li et al., 2021), and a handball training intervention for adults with MCI in a Chinese nursing home (Wei & Ji, 2014).

The two lower quality scoring studies not included in the meta-analysis evaluated physiotherapist-led interventions (Amjad et al. (2019); De Oliveira Silva et al. (2019)). The former involved aerobic training using an exercise bicycle three days per week for six weeks. Findings on cognitive outcomes favoured the intervention group (Mini-Mental State Examination, p= 0.032; Montreal Cognitive Assessment (p= 0.036), Trail Making Test-A (p=-0.005), and Trail Making Test-B (p=0.007). De Oliveira Silva and colleagues evaluated a three-month program of twice weekly group training sessions, including balance and aerobic exercises, strength training, and stretching. No differences were found in global cognition, only a significant difference between groups in the verbal fluency test (P = 0.05).

##### Summary

- In the meta-analysis of exercise studies (4 high quality) for which there was sufficient data, average effect size was 1.67 (95% CI 1.24-2.11) indicating a significant effect. Heterogeneity between studies was low (I^2 = 21.42%), though we found evidence for publication bias.
- A common characteristic of interventions where trial outcomes favoured the intervention was inclusion of a social element; all but one were delivered in groups.
- Interventions in successful trials were delivered by a range of facilitators and across different settings.

### 3.3 Multi-domain interventions (n=11)

#### 3.3.1 Studies rated as highest quality (n=2)

There was insufficient data available to include either of the higher quality studies in this category in our meta-analysis; both reported significant effects on cognition that favoured the intervention. Tao conducted a study evaluating a multi-domain intervention (Tao et al., 2019). It evaluated regular Baduanjin (traditional Chinese mind-body exercises) and health education, delivered by two professional coaches with over five years of teaching experience in Baduanjin practices, over 24 weeks. With an adherence rate of 82.6%, authors found a significant increase in cognition in the Baduanjin compared to non-exercise group (p = .050). Griffiths et al. (2020) tested a physical movement (up to 30 minutes) and cognitive stimulating (up to 90 minutes) activities intervention over 12 weeks, without using of technology and led by three occupational therapists. The intervention found improvements in attention (p = 0.018), memory (immediate, p = 0.001; delayed recall, p = 0.001). and executive function (p = 0.029) in the intervention group.

#### 3.3.2 Studies rated as lower quality (n=9)

We were able to include 5/9 of these studies in our meta-analysis, of which only two were associated with outcomes favouring the intervention. These evaluated individual, thrice weekly balance, cardio, strength and cognitively challenging activities facilitated by a physiotherapist with experience (Uysal et al., 2023) and a holistic, multicomponent health intervention group, involving strategies to manage cognitive loss and promotion of healthy lifestyle, led by a social worker that had worked for two years with neurocognitive conditions (Young et al., 2017). Three of the studies included in metanalysis did not find significant results (Yu et al., 2019; Lam et al., 2015; Law et al., 2019; See Table 2/Figure 4). Law et al. (2019), which found no within-group or between-group differences, did found significant differences in individual cognitive subdomains.

**Figure 4:**
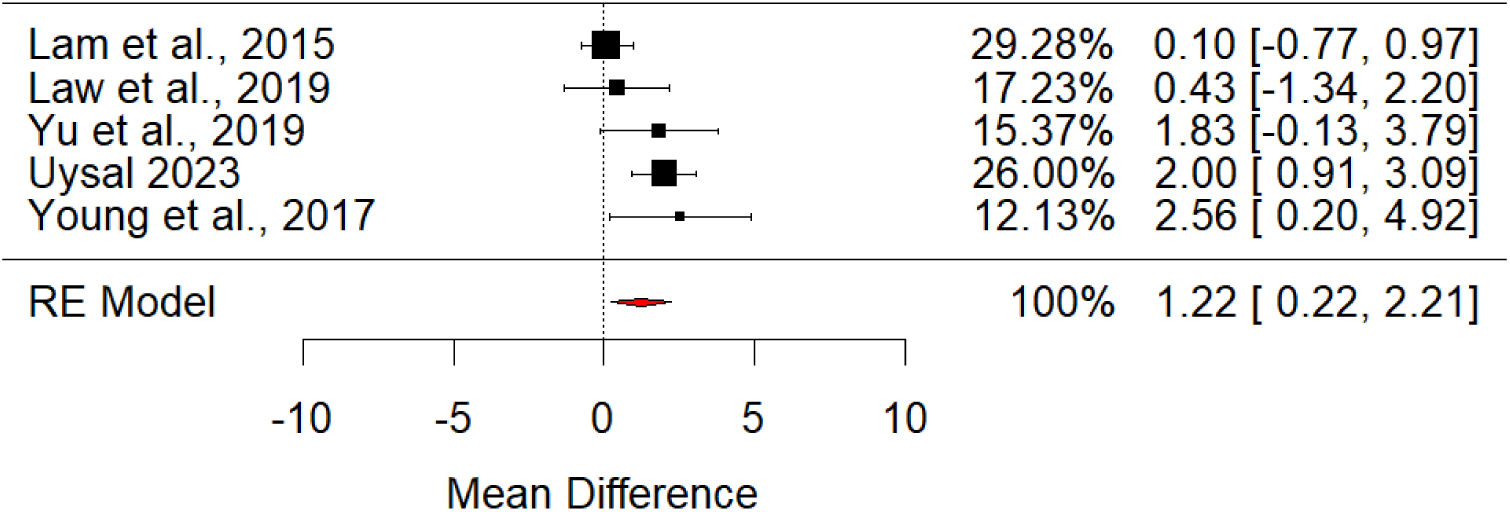
Meta analysis of multidomain interventions.

Of those excluded, from meta-analysis, three reported cognitive outcomes that favoured the intervention. Kwan et al. (2020) tested 12 weeks of an mhealth-delivered goal-setting and behavioural change intervention. Liu et al. (2023) tested a 9-month mindfulness meditation, cognitive training, physical exercise, and nutrition counselling group intervention. This last component was provided by a physician, while the group sessions were supervised by trained research assistants. They also included a homework component. This was the only intervention to include participants with subjective cognitive concerns. The study found a significant between-group difference favouring the intervention (change in overall cognition z score: 0.20, 95% CI: 0.05, 0.35). Chatterjee and colleagues (2022) conducted a 6-month trial on an intervention that compared: (A) computerised cognitive training (CBCT), (B) CBCT and Mediterranean diet, (C) CBCT, Mediterranean diet and physiotherapy. All intervention groups showed better cognition performance compared with the active control condition, with group C showing the greatest difference.

Finally, Horie and collaborators (2016) conducted a 12-month multi-domain intervention with moderate-intensity aerobic exercises or walking, a geriatrician visit and group nutritional counselling. There were no significant time-group interactions on cognitive tests, which were secondary outcomes.

##### Summary

- We conducted a metanalysis of 6/11 multidomain interventions for which there was sufficient data. The standardised effect size of 1.22 (0.22-2.21) indicated a significant overall effect. Heterogeneity between studies was moderate to substantial (I^2 = 57.30%). We found evidence for publication bias.
- No high-quality multicomponent intervention studies were included in the metanalysis; A study in China comparing mind-body exercises with a health promotion intervention, and a study in Thailand, which combined physical activity and cognitive stimulating activities without the use of technology.

### 3.5 Arts and creative expression (n=3)

All three arts and creation interventions took place in China and were in group settings. Two were rated as being of higher quality, of which one reported outcomes that favoured the intervention. Zhao et al. (2018) evaluated 25 sessions of creative expression program over 4 months facilitated by professional therapists. They found a significant difference in overall cognition (Montreal Cognitive Assessment; F=21.47, P,0.001), as well as other cognitive subdomains memory (Chinese Version of the Auditory Verbal Learning Test - immediate recall; F=4.81, P=0.023 and delay recall; F=3.98, P=0.012), language function (Chinese Version of the Category Verbal Fluency Test; F=3.91, P=0.017), among others. Luo and colleagues’ study (2023) evaluated six months of weekly sessions of story-telling and art making through group online classes and videoconferences with an instructor. Authors did find significant differences in various cognition outcomes (for instance: Montreal Cognitive Assessment, p = 0.012 and Mini-Mental State Examination, p = 0.035) between groups, but these were no longer significant after correcting using the False Discovery Rate. In a study not rated as being of higher quality, Lin et al. (2022) tested a similar intervention with verbal and non-verbal expression components, delivered face-to-face. They found significant differences in general cognition, favouring the intervention (mean difference =−0.905, 95% CI = −1.748 to −0.062, P = 0.038).

#### Summary

- One higher quality and one lower quality study found that face to face creative expression workshops led by professional therapists over six and four months were associated with improved cognitive outcomes, relative to control groups in China.
- Another face-to-face modality intervention that met 5/6 quality criteria found significant effects in cognition, but these were no longer significant after statistical correction.

Random Effects (RE) model combining effect size estimates on global cognition (standardised mean differences).

Funnel plot of the standardised mean difference in global cognition plotted against standard deviation.

## Discussion

This is to our knowledge the first systematic review of RCT evidence for non-pharmacological interventions in older adults with cognitive concerns to focus on LMIC. Overall, interventions reduced cognitive decline, which provides a positive message for the potential for dementia prevention. We found replicated evidence for exercise and multidomain interventions. There was insufficient data to meta-analyse the creative arts category. Most multidomain interventions included an exercise component, and several incorporated health education and/or cognitively challenging activities, healthy lifestyle education, carer-patient support, and creative activities.

Studies included in this review originated mostly from China, indicating a need for high-quality evidence from underrepresented regions, such as Africa, Latin America, and other Asian countries. This concerning global evidence gap is being addressed by Worldwide fingers, which seeks to implement the successful FINGERS intervention, adapting it for local contexts. LatAm-FINGERS is currently testing a multidomain intervention that includes an adapted version of dietary interventions previously implemented in higher income countries, for example finding interchangeable alternatives for food groups with similar nutritional value that can be available and accessible in countries such as Latino America (Crivelli et al., 2023). A consortium – Africa-FINGERS is planning similar implementation work across several Africa countries (*Network Groups - FBHI*, 2023).

Future interventions need to consider the fast-changing realities of different regions, with factors such as climate change impacting areas that could influence health outcomes (Wheeler & von Braun, 2013) such as food security. The frequency and intensity of interventions, and the need for highly trained or specialised facilitators, as well as monitoring strategies or other ways to ensure compliance, were discussed in several studies as relevant to determining whether implementation could be feasible outside research settings (e.g.

Amjad et al., 2019; Lin et al., 2022; De Oliveira Silva et al., 2019; Song & Yu, 2019). For example, while dietary changes have demonstrated positive effects on cognitive health before (Chen et al., 2019), focusing solely on this single behaviour may not represent a viable and comprehensive solution, especially depending on food availability (Kivipelto et al., 2018).

All the included studies recruited and randomised individual participants; we did not identify any cluster randomised controlled trials, though one is planned (MIND-China). Individualistic approaches to dementia prevention have been criticised (Leibing, 2018), with recent London-based studies concluding that structural inequalities associated with ethnic and socioeconomic status may contribute more to individual dementia risk than lifestyle choices (Bothongo et al., 2022). People with cognitive symptoms without dementia have described how burden of responsibility for managing dementia risk, without access to appropriate support, sometimes termed ‘responsibilisation’, can cause distress (Poppe et al., 2020). In coproduction of a multimodal intervention for people with mild memory concerns, a focus on addressing mental wellbeing and support to meet personalised goals emerged (Cooper et al., 2021). Such approaches were evaluated in lower quality multimodal studies and showed some promise (Uysal et al., 2023; Young et al., 2017). While almost all included trials reported a cognitive primary outcome, other outcomes, such as personalised goal attainment have been used in dementia non-pharmacological trials, as they can be more sensitive to, and perhaps more possible to change (Cooper et al., 2024). Some studies included other potentially relevant outcomes such as quality of life and activities of daily living. Future research could explore what outcomes matter most to people with cognitive concerns, and are more amenable to change, and measure lifestyle changes.

For comparability, we focused on global cognition, which were the most commonly identified primary outcomes across included studies, though measures of subdomains can be more sensitive in detecting subtle changes (Garg et al., 2022). Cognitive decline is a predictor but not the same as dementia diagnosis. Lack of evidence of efficacy is not evidence of inefficacy; our main finding in favour of group exercise as the most implementation-ready intervention could have been driven by the size of the evidence base. We did not find replicated evidence that any one type of intervention was ineffective.

There are inequalities in research participation, which are likely to be accentuated in LMIC where access to health care is more restricted (Cooper et al., 2014). We identified significant publication bias, indicating that inequality of access to open science and a robust evidence base may compound the multitude of other inequalities between global North and South. Emerging efforts to adapt and test preventive interventions will provide a much-needed boost in high-quality evidence from underrepresented regions. While individual approaches to prevention have been criticised in contexts where structural, socioeconomic inequalities, may be more effective in reducing dementia burden, for those who seek help for cognitive concerns, considering evidence for what works for local populations, using tailored approaches to avoid a “one size fits all” approach, and considering what outcomes matter most in this group may be helpful next steps. We plan such a programme in Chile, which we hope will inform future global approaches for managing cognitive concerns.

## Data Availability

All references used were obtained from online data bases.

## Supplementary material 1: URL to search strategy

Pubmed example:

(((((cognitive impairment) OR (cognitive decline)) OR (dementia)) AND ((((((RCT) OR (Trial)) OR (Intervention))) AND (((((((((((((((((((((((((((((((((((((((((((((((((((((((((((((((((((((((((((((((((((((((((((((((((((((((((((((((((((((((((((((((((((((Afghanistan) OR (Guinea)) OR (Korea)) OR (Burkina)) OR (Burundi)) OR (Liberia)) OR (Central African Republic)) OR (Madagascar)) OR (Chad) ) OR (Malawi)) OR (Congo)) OR (Mali)) OR (Eritrea)) OR (Mozambique)) OR (Ethiopia)) OR (Niger)) OR (Gambia)) OR (Rwanda)) OR (Guinea)) OR (Sierra Leone)) OR (Angola)) OR (India)) OR (Philippines)) OR (Algeria)) OR (Indonesia)) OR (Samoa)) OR (Bangladesh)) OR (Iran)) OR (São Tomé and Principe)) OR (Benin)) OR (Kenya)) OR (Senegal)) OR (Bhutan)) OR (Kiribati)) OR (Solomon Islands)) OR (Bolivia)) OR (Kyrgyz Republic)) OR (Sri Lanka)) OR (Cabo Verde)) OR (Lao)) OR (Tanzania)) OR (Cambodia)) OR (Lebanon)) OR (Tajikistan)) OR (Cameroon)) OR (Lesotho)) OR (Timor-Leste)) OR (Comoros)) OR (Mauritania)) OR (Tunisia)) OR (Congo)) OR (Micronesia)) OR (Ukraine)) OR (Côte d’Ivoire)) OR (Mongolia)) OR (Uzbekistan)) OR (Djibouti)) OR (Morocco)) OR (Vanuatu)) OR (Egypt)) OR (Myanmar)) OR (Vietnam)) OR (El Salvador)) OR (Nepal)) OR (West Bank and Gaza)) OR (Eswatini)) OR (Nicaragua)) OR (Zimbabwe)) OR (Ghana)) OR (Nigeria)) OR (Haiti)) OR (Pakistan)) OR (Honduras)) OR (Papua New Guinea)) OR (Albania)) OR (Fiji)) OR (Namibia)) OR (American Samoa)) OR (Gabon)) OR (North Macedonia)) OR (Argentina)) OR (Georgia)) OR (Palau)) OR (Armenia)) OR (Guatemala)) OR (Peru)) OR (Grenada)) OR (Paraguay)) OR (Azerbaijan)) OR (Belarus)) OR (Guyana)) OR (Russian Federation)) OR (Belize)) OR (Iraq)) OR (Serbia)) OR (Bosnia and Herzegovina)) OR (Jamaica)) OR (South Africa)) OR (Botswana)) OR (Jordan)) OR (St. Lucia)) OR (Brazil)) OR (Kazakhstan)) OR (St. Vincent and the Grenadines)) OR (Bulgaria)) OR (Kosovo)) OR (Suriname)) OR (China)) OR (Libya)) OR (Thailand)) OR (Colombia)) OR (Malaysia)) OR (Tonga)) OR (Costa Rica)) OR (Maldives)) OR (Türkiye)) OR (Cuba)) OR (Marshall Islands)) OR (Turkmenistan)) OR (Dominica)) OR (Mauritius)) OR (Tuvalu)) OR (Dominican Republic)) OR (Mexico)) OR (Equatorial Guinea)) OR (Mexico)) OR (Equatorial Guinea)) OR (Moldova)) OR (Ecuador)) OR (Montenegro)) OR (Costa Rica)) OR (Venezuela)) OR (Uruguay)) OR (Chile)) OR (China)) OR (Romania)) OR (Panama)) OR (Mauritius)) AND ((y_10[Filter]) AND (aged[Filter])) AND (effect*) Filters: Aged: 65+ years, Middle Aged: 45-64 years, from 2010/1/1 - 2023/1/1

Scielo example

Scielo (Includes all LATAM and south africa)

(Demencia) OR (Deterioro cognitivo) OR (Cogni*) OR (deterioro cognitivo level) OR (cognitive impairment) OR (cognitive decline) OR (Dementia) AND (Intervención) OR (Ensayo controlado aleatorizado) OR (RCT) OR (Trial)

FILTERED to exclude Spain

Lilacs (includes LATAM and central america)

(demencia) OR (deterioro cognitivo) OR (cogni*) OR (deterioro cognitivo leve) OR (cognitive impairment) OR (cognitive decline) OR (dementia) AND (intervención) OR (ensayo controlado aleatorizado) OR (rct) OR (trial) AND ( db:("LILACS") AND mj:("Enfermedad de Alzheimer" OR "Demencia" OR "Anciano" OR "Cognición" OR "Disfunción Cognitiva" OR "Pruebas Neuropsicológicas" OR "Demencia Vascular" OR "Salud del Anciano" OR "Memoria" OR "Neuropsicología" OR "Trastornos de la Memoria" OR "Demencia Frontotemporal") AND type_of_study:("clinical_trials" OR "evaluation_studies")) AND (year_cluster:[2010 TO 2023])

## References

Amjad, I., Toor, H., Niazi, I. K., Afzal, H., Jochumsen, M., Shafique, M., Allen, K., Haavik, H., & Ahmed, T. (2019). Therapeutic effects of aerobic exercise on EEG parameters and higher cognitive functions in mild cognitive impairment patients. International Journal of Neuroscience, 129(6), 551–562. 10.1080/00207454.2018.1551894

Bothongo, P. L. K., Jitlal, M., Parry, E., Waters, S., Foote, I. F., Watson, C. J., Cuzick, J., Giovannoni, G., Dobson, R., Noyce, A. J., Mukadam, N., Bestwick, J. P., & Marshall, C. R. (2022). Dementia risk in a diverse population: A single-region nested case-control study in the East End of London. The Lancet Regional Health - Europe, 15, 100321. 10.1016/j.lanepe.2022.100321

Chatterjee, P., Kumar, D. A., Naqushbandi, S., Chaudhary, P., Khenduja, P., Madan, S., … & Singh, V. (2022). Effect of Multimodal Intervention (computer based cognitive training, diet and exercise) in comparison to health awareness among older adults with Subjective Cognitive Impairment (MISCI-Trial)—A Pilot Randomized Control Trial. Plos one, 17(11), e0276986.

Chen, X., Maguire, B., Brodaty, H., & O’Leary, F. (2019). Dietary Patterns and Cognitive Health in Older Adults: A Systematic Review. Journal of Alzheimer’s Disease, 67(2), 583–619. 10.3233/JAD-180468

Cooper, C., Ketley, D., & Livingston, G. (2014). Systematic review and meta-analysis to estimate potential recruitment to dementia intervention studies. International Journal of Geriatric Psychiatry, 29(5), 515–525. 10.1002/gps.4034

Cooper, C., Mansour, H., Carter, C., Rapaport, P., Morgan-Trimmer, S., Marchant, N. L., Poppe, M., Higgs, P., Brierley, J., Solomon, N., Budgett, J., Bird, M., Walters, K., Barber, J., Wenborn, J., Lang, I. A., Huntley, J., Ritchie, K., Kales, H. C., … Palomo, M. (2021). Social connectedness and dementia prevention: Pilot of the APPLE-Tree video-call intervention during the Covid-19 pandemic. Dementia, 14713012211014382–14713012211014382. 10.1177/14713012211014382

Cooper, C., Vickerstaff, V., Barber, J., Phillips, R., Ogden, M., Walters, K., … & Budgett, J. (2024). A psychosocial goal-setting and manualised support intervention for independence in dementia (NIDUS-Family) versus goal setting and routine care: a single-masked, phase 3, superiority, randomised controlled trial. The Lancet Healthy Longevity, 5(2), e141–e151.

De Oliveira Silva, F., Ferreira, J. V., Plácido, J., Sant’Anna, P., Araújo, J., Marinho, V., Laks, J., & Camaz Deslandes, A. (2019). Three months of multimodal training contributes to mobility and executive function in elderly individuals with mild cognitive impairment, but not in those with Alzheimer’s disease: A randomized controlled trial. Maturitas, 126, 28–33. 10.1016/j.maturitas.2019.04.217

Deeks, J. J., Bossuyt, P. M., Leeflang, M. M., & Takwoingi, Y. (Eds.). (2023). Cochrane handbook for systematic reviews of diagnostic test accuracy. John Wiley & Sons.

Garg, D., Gupta, A., Agarwal, A., Mishra, B., Srivastava, M. V. P., Basheer, A., & Vishnu, V. Y. (2022). Latest Trends in Outcome Measures in Dementia and Mild Cognitive Impairment Trials. Brain Sciences, 12(7), Article 7. 10.3390/brainsci12070922

Geda, Y. E. (2012). Mild cognitive impairment in older adults. Current Psychiatry Reports, 14(4), 320–327. 10.1007/s11920-012-0291-x

Griffiths, J., Thaikruea, L., Wongpakaran, N., Munkhetvit, P., Kittisares, A., & Varnado, P. (2020). Effects of Combined Physical Movement Activity and Multifaceted Cognitive Training in Older People with Mild Neurocognitive Disorder in a Rural Community: A Randomized Control Trial. Dementia and Geriatric Cognitive Disorders, 49(2), 194–201. 10.1159/000507922

Hong, Q. N., Fàbregues, S., Bartlett, G., Boardman, F., Cargo, M., Dagenais, P., Gagnon, M.-P., Griffiths, F., Nicolau, B., O’Cathain, A., Rousseau, M.-C., Vedel, I., & Pluye, P. (2018). The Mixed Methods Appraisal Tool (MMAT) version 2018 for information professionals and researchers. Education for Information, 34(4), 285–291. 10.3233/EFI-180221

Horie, N. C., Serrao, V. T., Simon, S. S., Gascon, M. R. P., Dos Santos, A. X., Zambone, M. A., Del Bigio de Freitas, M. M., Cunha-Neto, E., Marques, E. L., Halpern, A., de Melo, M. E., Mancini, M. C., & Cercato, C. (2016). Cognitive Effects of Intentional Weight Loss in Elderly Obese Individuals With Mild Cognitive Impairment. The Journal of Clinical Endocrinology and Metabolism, 101(3), 1104–1112. 10.1210/jc.2015-2315

Horie, N. C., Serrao, V. T., Simon, S. S., Gascon, M. R. P., Dos Santos, A. X., Zambone, M. A., Del Bigio De Freitas, M. M., Cunha-Neto, E., Marques, E. L., Halpern, A., De Melo, M. E., Mancini, M. C., & Cercato, C. (2016). Cognitive Effects of Intentional Weight Loss in Elderly Obese Individuals With Mild Cognitive Impairment. The Journal of Clinical Endocrinology & Metabolism, 101(3), 1104–1112. 10.1210/jc.2015-2315

Kivipelto, M., Mangialasche, F., & Ngandu, T. (2018). Lifestyle interventions to prevent cognitive impairment, dementia and Alzheimer disease. Nature Reviews Neurology, 14(11), 653–666.

Klimova, B., & Maresova, P. (2017). Computer-Based Training Programs for Older People with Mild Cognitive Impairment and/or Dementia. Frontiers in Human Neuroscience, 11, 262. 10.3389/fnhum.2017.00262

Kwan, R. Y., Lee, D., Lee, P. H., Tse, M., Cheung, D. S., Thiamwong, L., & Choi, K.-S. (2020). Effects of an mHealth Brisk Walking Intervention on Increasing Physical Activity in Older People With Cognitive Frailty: Pilot Randomized Controlled Trial. JMIR mHealth and uHealth, 8(7), e16596. 10.2196/16596

Langoni, C. D. S., Resende, T. D. L., Barcellos, A. B., Cecchele, B., Knob, M. S., Silva, T. D. N., Da Rosa, J. N., Diogo, T. D. S., Filho, I. G. D. S., & Schwanke, C. H. A. (2019). Effect of Exercise on Cognition, Conditioning, Muscle Endurance, and Balance in Older Adults With Mild Cognitive Impairment: A Randomized Controlled Trial. Journal of Geriatric Physical Therapy, 42(2), E15–E22. 10.1519/JPT.0000000000000191

Leibing, A. (2018). Situated Prevention: Framing the “New Dementia”. *Journal of Law*, Medicine & Ethics, 46(3), 704–716. 10.1177/1073110518804232

Li, L., Liu, M., Zeng, H., & Pan, L. (2021). Multi-component exercise training improves the physical and cognitive function of the elderly with mild cognitive impairment: A six-month randomized controlled trial. Annals of Palliative Medicine, 10(8), 8919–8929. 10.21037/apm-21-1809

Lin, R., Luo, Y., Yan, Y., Huang, C., Chen, L., Chen, M., Lin, M., & Li, H. (2022). Effects of an art-based intervention in older adults with mild cognitive impairment: A randomised controlled trial. Age and Ageing, 51(7), afac144. 10.1093/ageing/afac144

Liu, X., Ma, Z., Zhu, X., Zheng, Z., Li, J., Fu, J., Shao, Q., Han, X., Wang, X., Wang, Z., Yin, Z., Qiu, C., & Li, J. (2023). Cognitive Benefit of a Multidomain Intervention for Older Adults at Risk of Cognitive Decline: A Cluster-Randomized Controlled Trial. The American Journal of Geriatric Psychiatry : Official Journal of the American Association for Geriatric Psychiatry, 31(3), 197–209. 10.1016/j.jagp.2022.10.006

Lü, J., Sun, M., Liang, L., Feng, Y., Pan, X., & Liu, Y. (2016). Effects of momentum-based dumbbell training on cognitive function in older adults with mild cognitive impairment: A pilot randomized controlled trial. Clinical Interventions in Aging, 11, 9–16. 10.2147/CIA.S96042

Luo, Y., Lin, R., Yan, Y., Su, J., Lin, S., Ma, M., & Li, H. (2023). Effects of Remote Expressive Arts Program in Older Adults with Mild Cognitive Impairment: A Randomized Controlled Trial. Journal of Alzheimer’s Disease, 91(2), 815–831. 10.3233/JAD-215685

Martin, A. M. (2006). Dementia: Key factors in recognition and support. Practice Nursing, 17(1), 12–16.

McGrattan, A. M., Zhu, Y., Richardson, C. D., Mohan, D., Soh, Y. C., Sajjad, A., … & Stephan, B. (2021). Prevalence and risk of mild cognitive impairment in low and middle-income countries: a systematic review. Journal of Alzheimer’s Disease, 79(2), 743–762.

McGrattan, A., van Aller, C., Narytnyk, A., Reidpath, D., Keage, H., Mohan, D., Su, T. T., Stephan, B., Robinson, L., & Siervo, M. (2022). Nutritional interventions for the prevention of cognitive impairment and dementia in developing economies in East-Asia: A systematic review and meta-analysis. Critical Reviews in Food Science and Nutrition, 62(7), 1838–1855. 10.1080/10408398.2020.1848785

Naheed, A., Hakim, M., Islam, M. S., Islam, M. B., Tang, E. Y. H., Prodhan, A. A., Amin, M. R., Stephan, B. C. M., & Mohammad, Q. D. (2023). Prevalence of dementia among older age people and variation across different sociodemographic characteristics: A cross-sectional study in Bangladesh. The Lancet Regional Health - Southeast Asia, 17. 10.1016/j.lansea.2023.100257

Network groups—FBHI. (2023, July 15). https://fbhi.se/world-wide-fingers-collaboration-groups/

Ngandu, T., Lehtisalo, J., Korkki, S., Solomon, A., Coley, N., Antikainen, R., Bäckman, L., Hänninen, T., Lindström, J., Laatikainen, T., Paajanen, T., Havulinna, S., Peltonen, M., Neely, A. S., Strandberg, T., Tuomilehto, J., Soininen, H., & Kivipelto, M. (2022). The effect of adherence on cognition in a multidomain lifestyle intervention (FINGER). Alzheimer’s & Dementia, 18(7), 1325–1334. 10.1002/alz.12492

Ngandu, T., Lehtisalo, J., Solomon, A., Levälahti, E., Ahtiluoto, S., Antikainen, R., Bäckman, L., Hänninen, T., Jula, A., Laatikainen, T., Lindström, J., Mangialasche, F., Paajanen, T., Pajala, S., Peltonen, M., Rauramaa, R., Stigsdotter-Neely, A., Strandberg, T., Tuomilehto, J., … Kivipelto, M. (2015). A 2 year multidomain intervention of diet, exercise, cognitive training, and vascular risk monitoring versus control to prevent cognitive decline in at-risk elderly people (FINGER): A randomised controlled trial. The Lancet, 385(9984), 2255–2263. 10.1016/S0140-6736(15)60461-5

Palafox, B., McKee, M., Balabanova, D., AlHabib, K. F., Avezum, A. J., Bahonar, A., Ismail, N., Chifamba, J., Chow, C. K., Corsi, D. J., Dagenais, G. R., Diaz, R., Gupta, R., Iqbal, R., Kaur, M., Khatib, R., Kruger, A., Kruger, I. M., Lanas, F., … Yusuf, S. (2016). Wealth and cardiovascular health: A cross-sectional study of wealth-related inequalities in the awareness, treatment and control of hypertension in high-, middle- and low-income countries. International Journal for Equity in Health, 15(1), 199. 10.1186/s12939-016-0478-6

Parra, M. A., Butler, S., McGeown, W. J., Brown Nicholls, L. A., & Robertson, D. J. (2019). Globalising strategies to meet global challenges: The case of ageing and dementia. Journal of Global Health, 9(2), 020310. 10.7189/jogh.09.020310

Posit team (2023). RStudio: Integrated Development Environment for R. Posit Software, PBC, Boston, MA. URL http://www.posit.co/.

Prince, M., Acosta, D., Albanese, E., Arizaga, R., Ferri, C. P., Guerra, M., Huang, Y., Jacob, K., Jimenez-Velazquez, I. Z., Rodriguez, J. L., Salas, A., Sosa, A. L., Sousa, R., Uwakwe, R., van der Poel, R., Williams, J., & Wortmann, M. (2008). Ageing and dementia in low and middle income countries–Using research to engage with public and policy makers. International Review of Psychiatry, 20(4), 332–343. 10.1080/09540260802094712

Rosenberg, A., Mangialasche, F., Ngandu, T., Solomon, A., & Kivipelto, M. (2020). Multidomain Interventions to Prevent Cognitive Impairment, Alzheimer’s Disease, and Dementia: From FINGER to World-Wide FINGERS. J Prev Alzheimers Dis, 7(1), 29–36. MEDLINE. 10.14283/jpad.2019.41

Song, D., & Yu, D. S. F. (2019). Effects of a moderate-intensity aerobic exercise programme on the cognitive function and quality of life of community-dwelling elderly people with mild cognitive impairment: A randomised controlled trial. International Journal of Nursing Studies, 93, 97–105. 10.1016/j.ijnurstu.2019.02.019

Sungkarat, S., Boripuntakul, S., Kumfu, S., Lord, S. R., & Chattipakorn, N. (2018). Tai Chi Improves Cognition and Plasma BDNF in Older Adults With Mild Cognitive Impairment: A Randomized Controlled Trial. Neurorehabilitation and Neural Repair, 32(2), 142–149. 10.1177/1545968317753682

Tao, J., Liu, J., Chen, X., Xia, R., Li, M., Huang, M., Li, S., Park, J., Wilson, G., Lang, C., Xie, G., Zhang, B., Zheng, G., Chen, L., & Kong, J. (2019). Mind-body exercise improves cognitive function and modulates the function and structure of the hippocampus and anterior cingulate cortex in patients with mild cognitive impairment. NeuroImage: Clinical, 23, 101834. 10.1016/j.nicl.2019.101834

Uysal, İ., Başar, S., Aysel, S., Kalafat, D., & Büyüksünnetçi, A. Ö. (2023). Aerobic exercise and dual-task training combination is the best combination for improving cognitive status, mobility and physical performance in older adults with mild cognitive impairment. Aging Clinical and Experimental Research, 35(2), 271–281. 10.1007/s40520-022-02321-7

Walker, R., & Paddick, S.-M. (2019). Dementia prevention in low-income and middle-income countries: A cautious step forward. The Lancet Global Health, 7(5), e538–e539. 10.1016/S2214-109X(19)30169-X

Wang, J., Xie, J., Li, M., Ren, D., Li, Y., He, Y., Ao, Y., & Liao, S. (2022). Finger exercise alleviates mild cognitive impairment of older persons: A community-based randomized trial. Geriatric Nursing, 47, 42–46. 10.1016/j.gerinurse.2022.06.014

Wang, L., Wu, B., Tao, H., Chai, N., Zhao, X., Zhen, X., & Zhou, X. (2020). Effects and mediating mechanisms of a structured limbs-exercise program on general cognitive function in older adults with mild cognitive impairment: A randomized controlled trial. International Journal of Nursing Studies, 110, 103706. 10.1016/j.ijnurstu.2020.103706

Wei, X., & Ji, L. (2014). Effect of handball training on cognitive ability in elderly with mild cognitive impairment. Neuroscience Letters, 566, 98–101. 10.1016/j.neulet.2014.02.035

Wheeler, T., & von Braun, J. (2013). Climate Change Impacts on Global Food Security. Science, 341(6145), 508–513. 10.1126/science.1239402

Whitty, E., Mansour, H., Elisa Aguirre, Elisa Aguirre, Aguirre, E., Palomo, M., Charlesworth, G., Ramjee, S., Poppe, M., Brodaty, H., Kales, H. C., Morgan-Trimmer, S., Nyman, S. R., Lang, I. A., Lang, I., Walters, K., Petersen, I., Wenborn, J., Minihane, A. M., … Cooper, C. (2020). Efficacy of lifestyle and psychosocial interventions in reducing cognitive decline in older people: Systematic review. Ageing Research Reviews, 62, 101113. 10.1016/j.arr.2020.101113

Young, D. K., Ng, P. Y., Kwok, T., & Cheng, D. (2017). The effects of holistic health group interventions on improving the cognitive ability of persons with mild cognitive impairment: A randomized controlled trial. *Clinical Interventions in Aging*, Volume 12, 1543–1552. 10.2147/CIA.S142109

Zhao, J., Li, H., Lin, R., Wei, Y., & Yang, A. (2018). Effects of creative expression therapy for older adults with mild cognitive impairment at risk of Alzheimer’s disease: A randomized controlled clinical trial. *Clinical Interventions in Aging*, Volume 13, 1313– 1320. 10.2147/CIA.S161861

Zhu, Y., Wu, H., Qi, M., Wang, S., Zhang, Q., Zhou, L., Wang, S., Wang, W., Wu, T., Xiao, M., Yang, S., Chen, H., Zhang, L., Zhang, K. C., Ma, J., & Wang, T. (2018). Effects of a specially designed aerobic dance routine on mild cognitive impairment. Clinical Interventions in Aging, 13, 1691–1700. 10.2147/CIA.S163067

